# Automatic Measurements of Smooth Pursuit Eye Movements by Video-Oculography and Deep Learning-Based Object Detection

**DOI:** 10.1101/2020.11.09.20227736

**Authors:** Masakazu Hirota, Takao Hayashi, Emiko Watanabe, Atsushi Mizota

**Affiliations:** Department of Orthoptics, Faculty of Medical Technology, Teikyo University, Itabashi, Tokyo, Japan; Department of Ophthalmology, School of Medicine, Teikyo University, Itabashi, Tokyo, Japan

**Author notes:** **Corresponding author:** Masakazu Hirota, Address: 2-11-1 Kaga, Itabashi, Tokyo, 173-8605 Japan.

## Abstract

**Purpose:** The purpose of this study was to develop a technique combining video oculography (VOG) with single shot multibox detector (SSD) to accurately and quantitatively examine eye movements.

**Methods:** Eleven healthy volunteers (21.3 ± 0.9 years) participated in this study. Eye movements were recorded while tracking a target using a custom-made eye tracker. The subjects were asked to fixate their focus on the nose of the rabbit-like target (visual angle was 0.1°), which was manually moved to a distance of 1 meter by the examiner during the eye movement test. The test produced 500 images from the VOG external camera and these images were divided into 3 groups (300, 100, and 100) for training, verification, and testing. The performance of the SSD was evaluated with 75% average precision (AP_75_), and the relationship between the location of the fixated target (calculated by the SSD) and the positions of both eyes (recorded by the VOG) was analyzed.

**Results:** The AP_75_ of the SSD on one class of targets was 97.7%. The horizontal and vertical target locations significantly and positively correlated with the horizontal and vertical both eye positions (*adjusted R*^*2*^ ≥ 0.955, *P* < 0.001).

**Conclusions:** Our findings suggest that VOG with SSD is suitable for the evaluation of eye version movements in standard clinical assessments.

**Translational Relevance:** The combination of VOG and SSD can be used to evaluate the SPEM, and this method can be translated into clinical settings without changing the testing methods.

## Introduction

Strabismus is an eye movement disorder, and it is important to detect limitation of eye movement by confirming the version eye movement with smooth pursuit eye movement (SPEM)^1-8^ in nine directions.^9-11^ In most ophthalmology clinics, the examiner visually evaluates SPEM.^12, 13^ The examiner detects abnormalities in eye movements by carefully examining SPEM.^12, 13^ However, the current method does not allow recording them during the examination and does not quantitively evaluate the SPEM.

In the laboratory, several methods have been used to quantify eye movements, for example, the search coil method,^14, 15^ electrooculography,^16^ and video-oculography (VOG).^15^ These techniques can record eye movements of both eyes simultaneously while the subjects are have fixated their focus on a moving target. In particular, VOG is used to evaluate eye movements in strabismus patients with wide range of age groups from children to adults ^17-20^ because it can measure eye movements non-invasively and easily. However, laboratory methods have not been introduced in clinical practice for the following two basic reasons: first, the type of target is restricted. To achieve accuracy at the laboratory level, it is necessary to present a predetermined target to the subjects according to the programming codes. In children, the examiner uses a target, such as an anime or game character, to help keep their attention.^10, 18, 2-23^ Second, the VOG is operated with the display, which limits the flexibility of the examination. The target only makes a fixed movement, and the movement cannot be changed flexibly depending on the eye movement disorder.

We hypothesized that a combination of VOG and a deep learning-based object detection algorithm is one method of translating laboratory methods to the clinic. Deep learning-based object detection technology can predict the location and types of objects in one image.^24-26^ Furthermore, the algorithm of deep learning-based object detection can detect the objects in real-time in the movie with a processing speed of >30 fps. This processing speed is faster than that of conventional simpler algorithms that use raster scans per image.^27^ Therefore, we expected that by using VOG to record SPEM and deep learning-based object detection to record movements of the target, the combination system can measure the SPEM and target simultaneously without changing the clinical examination.

Thus, this study aimed to develop a technique that would combine VOG with deep learning-based object detection that can quantify SPEM. We evaluated the target location and both eye positions in healthy volunteers without changing the clinical method. The experiments were designed to be performed as close to the ophthalmological clinical setting as possible.

## Methods

### Subjects

Eleven volunteers (age, 21.3 ± 0.9 years [mean ± standard deviation]) participated in this study. All subjects underwent complete ophthalmologic examinations, including determination of ocular dominance using the hole-in-card test, best-corrected visual acuity at distance (5.0 m), near point of convergence, stereoscopic acuity (Titmus stereotest; Stereo Optical Co., Inc., Chicago, USA) at 40 cm, and heterophoria by alternate cover test both at near (33 cm) and at distance (5.0 m), and fundus examinations. Stereoacuity was converted to the logarithm of the arcsecond (log arcsec). Participants were excluded if they had a refractive error >±10.0 D.

Informed consent was obtained from all subjects after the nature and possible complications of the study were explained to them. This investigation adhered to the tenets of the World Medical Association Declaration of Helsinki. Furthermore, informed consent for publication of identifying information/images in an online open-access publication was obtained from a part of subjects. The Institutional Review Board of Teikyo University approved the experimental protocol and consent procedures (approval no. 18–161).

### Eye movement recordings

Eye movements were recorded during the tracking of the target using a custom-made eye tracker based on EMR-9 (NAC Image Technology Inc., Tokyo, Japan) (Fig. 1). The EMR-9 tracker determined the eye positions by detecting the corneal reflex and pupil center created by the reflection of a near-infrared light that can be adjusted by the half-mirror. The sampling rate was 240 Hz, the measurement angle was ±31° from the center of the scene camera, and the measurement error was 0.2°–0.5° (interquartile range) at a distance of 1.0 m. The EMR-9 controller merged the gaze of both eyes to the real scenes (resolution, 640 × 480 pixels) that were recorded by a scene camera with a sampling rate of 29.97 Hz and a delay of ≤52 ms.

**Figure 1.**
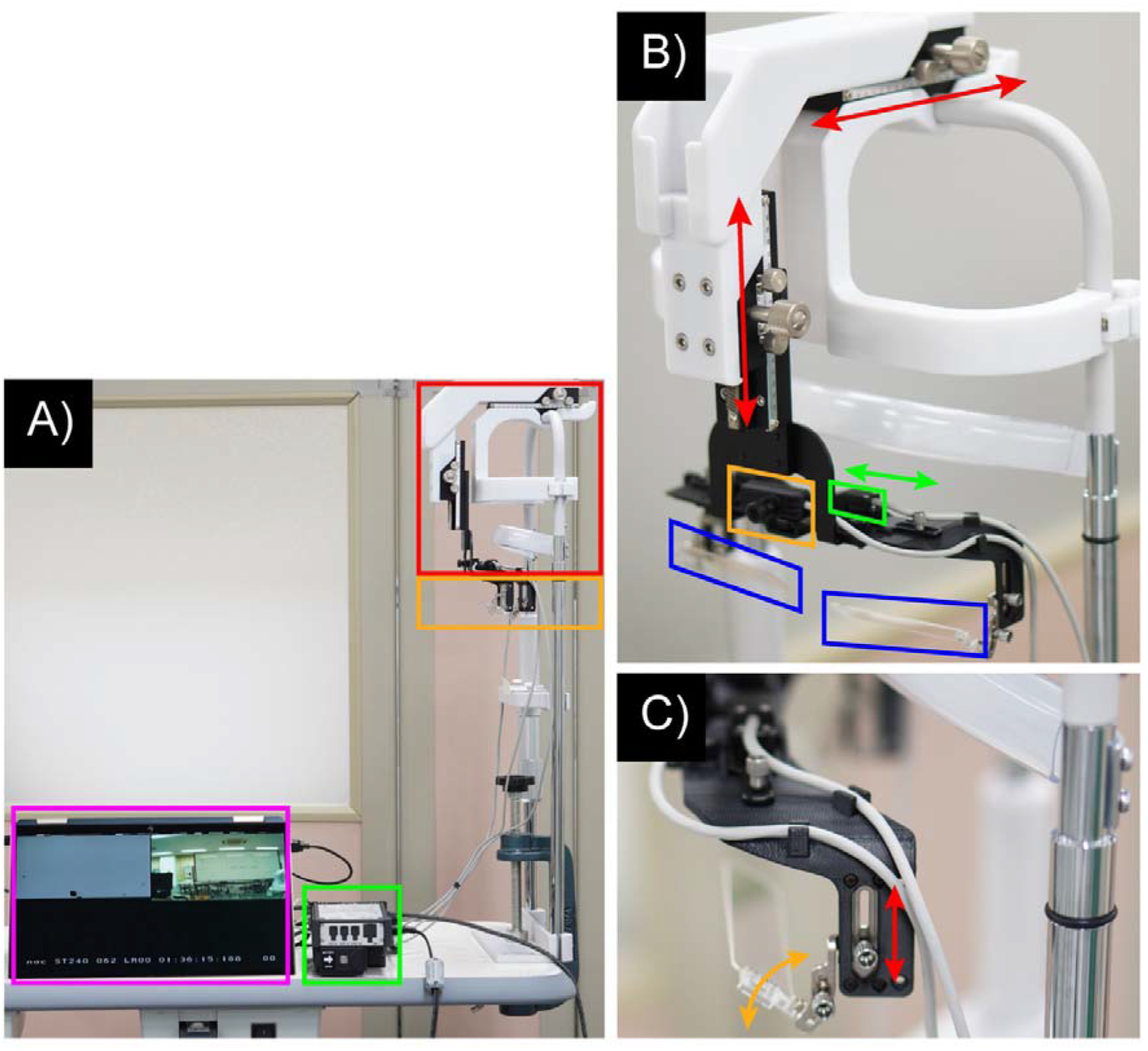
Custom-made eye tracker based on EMR-9. The exterior of the custom-made eye tracker was based on EMR-9 (A). The total area of the red and orange squares indicates EMR-9, and the red and orange squares indicate (B) and (C). The green square indicates the controller of the EMR-9. The purple square indicates the output image by EMR-9. In (B), the orange, green, and blue squares indicate the scene camera, eye camera (there is also one for the right eye on the other side), and half-mirrors, respectively. The scene camera is able to rotate 60°in pitch. Both eye cameras can horizontally shift to 1.3 cm (total, 2.6 cm) to adjust the pupillary distance (green two-direction arrow line). The base position of the EMR-9 can adjust 8.0 cm horizontally and vertically (red two-direction arrow lines). In (C), the half-mirror can shift to 2.5 cm vertically (red two-direction arrow line) and rotate 30° in pitch (orange two-direction arrow line).

All subjects underwent a calibration test under binocular conditions with the fully corrected glasses at 1.0 m before the eye movement test. Noise caused by lens reflection was noted in 2 of 11 participants and avoided by manually changing the tilt of the half-mirror. During the calibration, all subjects were asked to fixate a black nine-point target (visual angle, 0.1°) on a whiteboard. The center of the screen was defined as 0°, the right and upper halves of the screen were defined as the positive sides, and the left and lower halves were defined as the negative sides.

### Pilot Study

The accuracy of VOG was evaluated in an ideal environment as a pilot study. Two subjects participated in the pilot study. The target was a rabbit-like character (Fig. 2). The size of the target was 10 × 10 cm, which subtended a visual angle of 5.7° at 1.0 m. The target was displayed on a 24-inch liquid crystal monitor. The center of the monitor was defined as 0°, the right and upper halves of the monitor were defined as the positive sides, and the left and lower halves were defined as the negative sides. The target moved to ±10° with a random velocity of ≤10°/s. The subjects were seated in a well-lit room (600 lx) wearing their fully corrective spectacles. The subject’s head was fixed with a chin and forehead rest. The subjects were asked to fixate their focus on the nose of the target whose visual angle was 0.1° at 1.0 m for 60 s.

**Figure 2.**
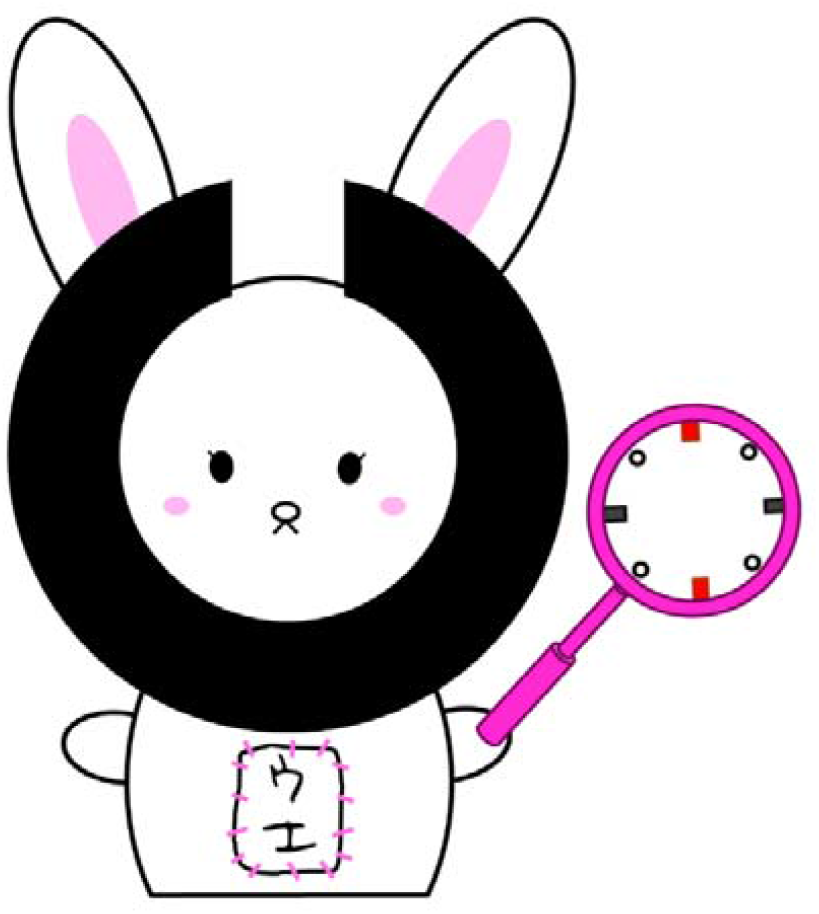
Examination target. The rabbit-like character is a mascot of the Department of Orthoptics, Teikyo University (created by Mika Suda). The subjects were asked to fixate on the nose of the target whose visual angle was 0.1° at 1.0 m during the eye movement tests.

### Eye movement test

The main study used the same target with pilot study (Fig 2). The target was manually moved within ±15° for 60 s by an examiner. All subjects were seated in a well-lit room (600 lx) wearing their fully corrective spectacles. The subject’s head was fixed with a chin rest and forehead rest. The subjects were asked to fixate on the nose of the target whose visual angle was 0.1° at 1.0 m during the eye movement test.

### Training, validation, and test sets

A total of 500 images were extracted from the video recordings of the eye movement test, which had been recorded by an EMR-9 scene camera for Subject 1. The training, validation, and test datasets were randomly divided into 300 (60%), 100 (20%), and 100 (20%) images, respectively.

### Object detection algorithm

An SSD was used.^25^ The SSD quantifies the output space of the bounding box into a set of default boxes regardless of the aspect ratio and scale for each feature map position. The network generates a score for the presence of each object category in each default box and makes adjustments to the box to better match the shape of the object in the prediction. The network combines the predictions from multiple feature maps with different resolutions to handle objects of different sizes.

The SSD was divided into the following two parts: shared feedforward convolutional network and set of subnetworks for classification and regression, which do not share computations. A pre-trained VGG16 convolutional neural network was used as a base network.^28, 29^ The target images were resized to 300 × 300 pixels in the preprocessing; then, data augmentation was applied to these target images. The data that were convolved 10 times were extracted as Source 1 (channel number, 512; feature size, 38 × 38) by normalizing the size. The data that were convolved 15 times were defined as Source 2 (channel number, 512; feature size, 19 × 19). The VGG output of Source 2 was entered into the extra module. The extra module convoluted Source 2 eight times, and the sources from 3 to 6 (feature sizes, 10 × 10, 5 × 5, 3 × 3, and 1 × 1, respectively) were output each time it was convoluted twice. Next, the sources were entered into the location and confidence module, and these data were convoluted once. The location module outputted the offset of the default bounding box. The confidence module outputted the confidence of each class for the default bounding box.

In the training phase, we set the following parameters: 100 epochs, batch size of 32, and Adam optimizer with the learning rate of 0.001. The validation was performed every 10 epochs.

The SSD model was evaluated by the average precision of the test dataset. The AP was calculated by the integral of precision and recall. The intersection of union was calculated by dividing the area of overlap between the predicted bounding box and the ground-truth bounding box that was included target name and target location manually defined by an examiner (MH) and the area of union of both bounding boxes. We defined an IoU ≥ 75% as “correct.” Subsequently, the results of the test dataset were classified as follows: true positive (TP), predicted bounding box has been covered with ground-truth bounding box (IoU ≥ 75%); false positive (FP), predicted bounding box has been covered with ground-truth bounding box (IoU < 75% and IoU ≠ 0); false negative (FN), predicted bounding box has not been covered with ground-truth bounding box (IoU = 0); and true negative, predicted bounding box and ground-truth bounding box did not exist. Precision was defined as the percentage at which the IoU can be correctly predicted with an accuracy of ≥75%. This was calculated by TP / (TP + FP). Recall was defined as the percentage at which the ratio that bounding box at apposition close to the correct answer can be predicted if the IoU is ≥75% and was calculated by TP / (TP + FN). The AP was calculated by the integral of precision and recall.

We used the software Python 3.6.5 on Windows 10 (Microsoft Co., Ltd., Redmond, WA, USA) with the following libraries: Matplotlib 3.3.2, Numpy 1.18.5, OpenCV 3.3.1, Pandas 1.1.3, Pytorch 1.6.0, Scikit-learn 0.23.2, and Seaborn 0.11.0.

### Calculating target location

The SSD drew the location of the object as the bounding box. The bounding box was computed from two coordinates of X_min_, Y_min_, and X_max_, Y_max_. The center of the object coordinates (Cx, Cy) was determined by ((X_max_ + X_min_) / 2, (Y_max_ + Y_min_) / 2). Thus, the target location was defined as the center of the bounding box, and a program that outputs the target location in each frame with the recording time synchronized with the EMR-9 findings to an Excel file (Microsoft Co., Ltd.) in the inference phase was implemented.

### Data analyses

Data on eye positions and pupil sizes of both eyes were exported to a CSV file. Data were excluded if the pupil diameter changed by >2 mm/frame due to blinking.^30^ Data were also excluded if the pupil diameter changed by >0.2 mm/frame over an average of 11 points and median of 5 points due to noise. A linearly interpolated value replaced the missing values. Horizontal and vertical eye movements were analyzed, and the SPEM and SEM were identified using a velocity-threshold identification (I-VT) filter.^31^ The I-VT filter was used to classify eye movements based on the velocity of the directional shifts of the eye. The saccade was defined as the median velocity of three consecutive windows >100°/s. Then, the eye position data at 240 Hz were synchronized with the target data at 29.97 Hz.

The waveforms of the target locations and eye positions peaked when the moving direction was reversed (Fig. 3). The latency of SPEM was calculated from the difference between the horizontal and vertical target location peaks and the dominant and nondominant eye position peaks. The latency of both eyes was calculated three times or more and averaged for each subject.

**Figure 3.**
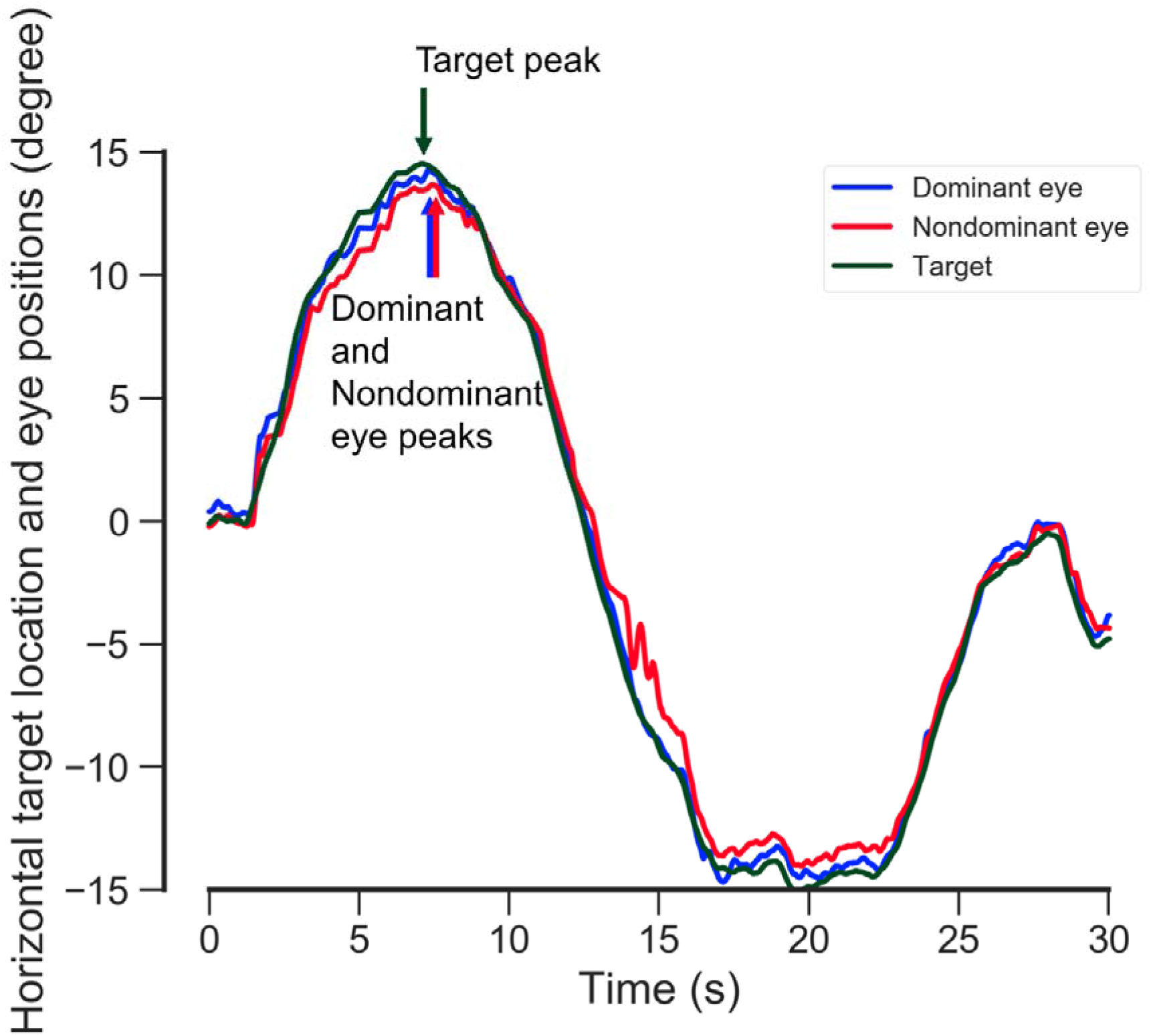
Graph showing the latencies of SPEM for the dominant and nondominant eyes. The blue, red, and green lines indicate the horizontal dominant and nondominant eye positions and target location during the eye movement test. The latencies of SPEM were calculated from the difference between the target location peak and the dominant and nondominant eye position peaks.

### Statistical analyses

The relationships between the target location and both eye positions were assessed using simple linear regression analysis. The difference between the latencies of the dominant and nondominant eyes was analyzed by paired *t*-tests. The relationship between latencies of horizontal and vertical SPEM within both eyes was assessed using single linear regression analysis.

The IBM SPSS Statistics version 26 software (IBM Corp., Armonk, NY, USA) was used to determine the significance of the differences, and a *P*-value < 0.05 was considered statistically significant.

## Results

### Pilot Study

#### Accuracy of VOG in conventional laboratory method

The target location was significantly correlated with both eye positions. The horizontal and vertical target locations were significantly and positively correlated with the horizontal dominant (*adjusted R*^*2*^ = 0.989, *P* < 0.001) and nondominant (*adjusted R*^*2*^ = 0.989, *P* < 0.001) eye positions. The vertical dominant (*adjusted R*^*2*^ = 0.987, *P* < 0.001) and nondominant (*adjusted R*^*2*^ = 0.987, *P* < 0.001) eye positions are shown in Figure 4.

**Figure 4.**
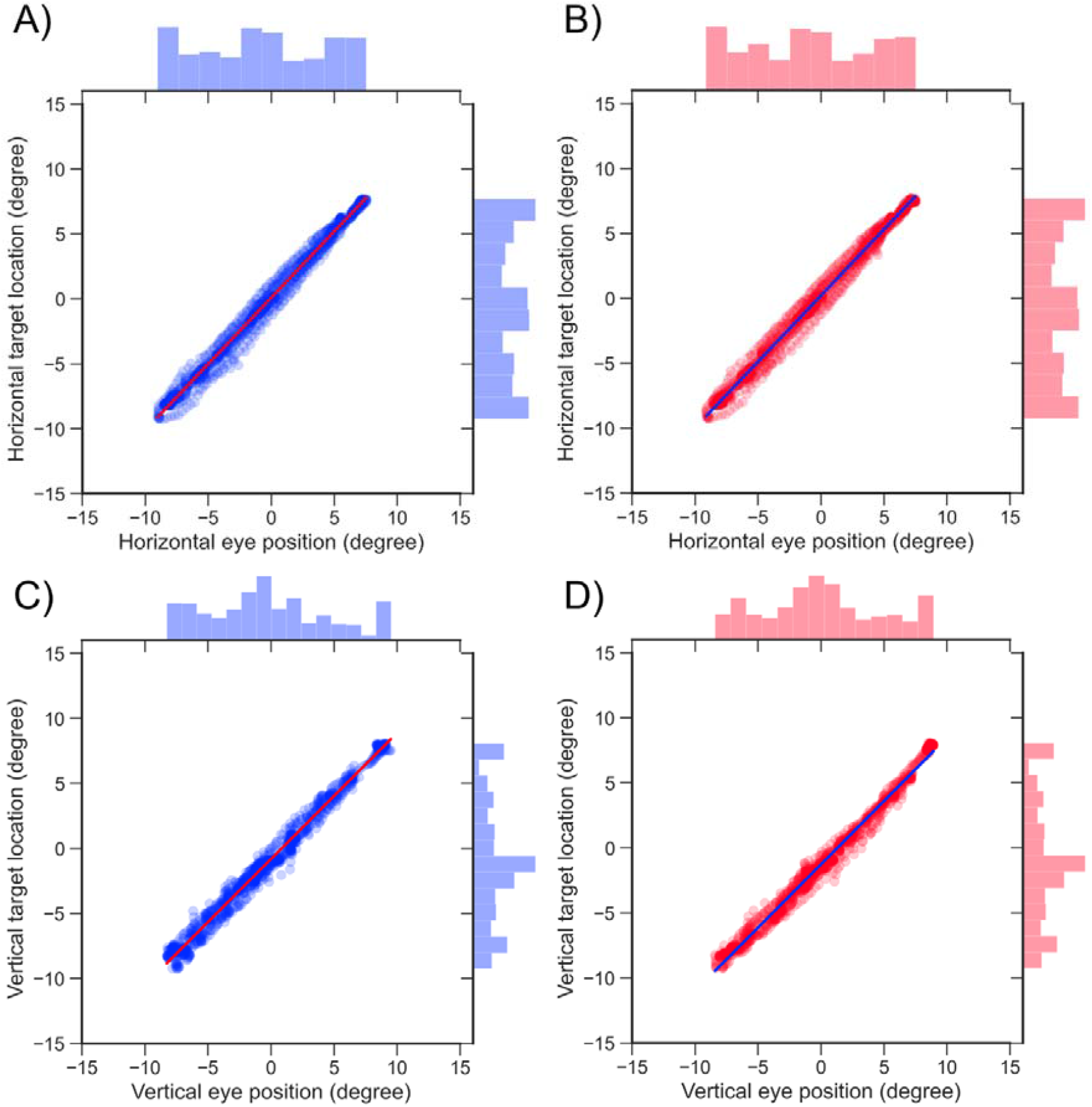
Correlations between horizontal (A, B) and vertical (C, D) target locations and eye positions in a pilot study (n = 2) The blue and red dots indicate the relationships between the target and dominant or nondominant eye velocity. The red and blue lines indicate the regression lines. The histograms at the upper and right sides indicate the distribution between the target location and the dominant or nondominant eye position.

### Main study

#### Demographics

The demographics of the subjects are presented in Table 1. The mean ± standard deviation of the refractive errors (spherical equivalents) of the dominant eye was −2.95 ± 2.46 diopters (D), and that of the nondominant eye was −2.70 ± 2.60 D. The best-corrected visual acuity was 0.0 logMAR units or better in all subjects. The average heterophoria was −1.3 ± 0.9 prism diopters (PD) at distance and −3.1 ± 4.4 PDs at near. All healthy individuals had a stereo acuity of 1.60 log arcsec.

#### Integration of deep learning-based object detection and VOG

A representative result for our single shot multibox detector (SSD) is shown in Movie 1 and Figure 5. The SSD could correctly track a target that an examiner manually moved. The average precision of the SSD on one class of targets was 97.7% in the test dataset.

**Figure 5.**
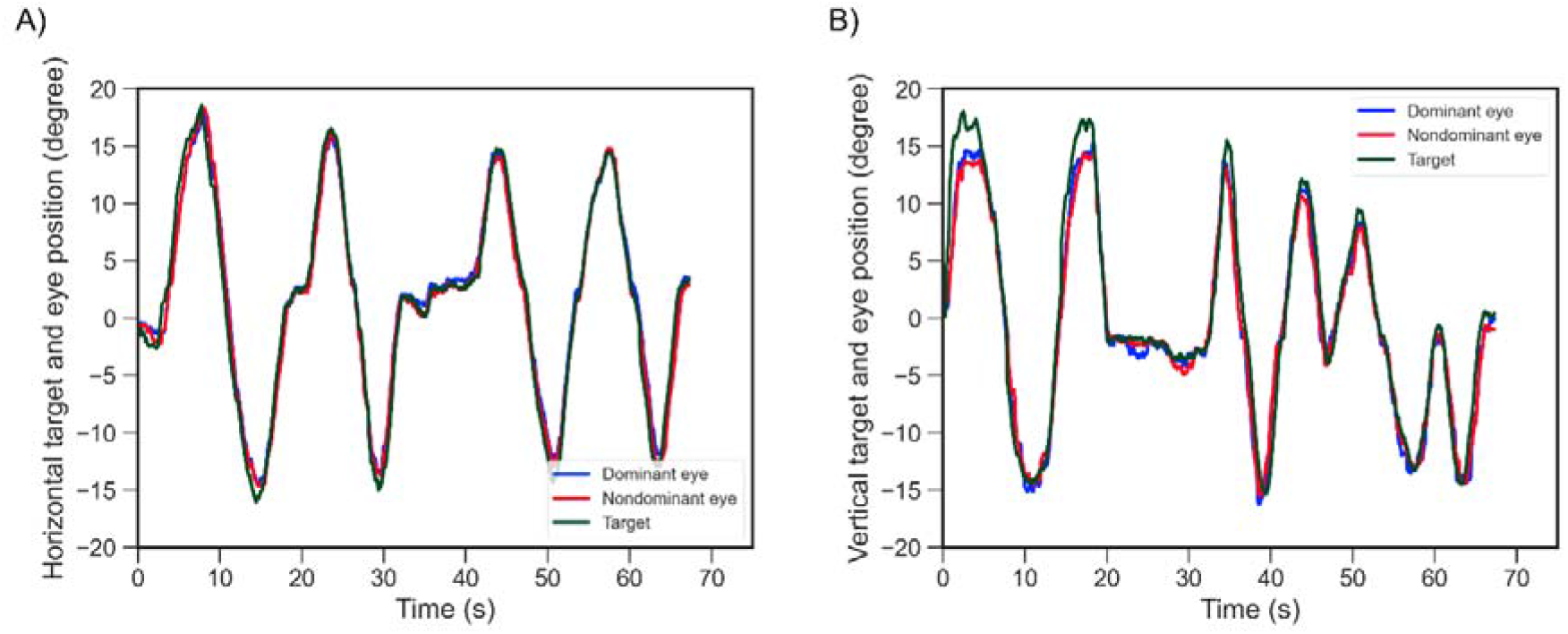
Horizontal (A) and vertical (B) target locations and eye positions when the examiner moved the target by hand. The blue, red, and green lines indicate the horizontal dominant and nondominant eye positions and target location during the eye movement test.

The horizontal and vertical target locations were significantly and positively correlated with the horizontal dominant (*adjusted R*^*2*^ = 0.984, *P* < 0.001) and nondominant (*adjusted R*^*2*^ = 0.983, *P* < 0.001) eye positions. The vertical dominant (*adjusted R*^*2*^ = 0.955, *P* < 0.001) and nondominant (*adjusted R*^*2*^ = 0.964, *P* < 0.001) eye positions are shown in Figure 6.

**Figure 6.**
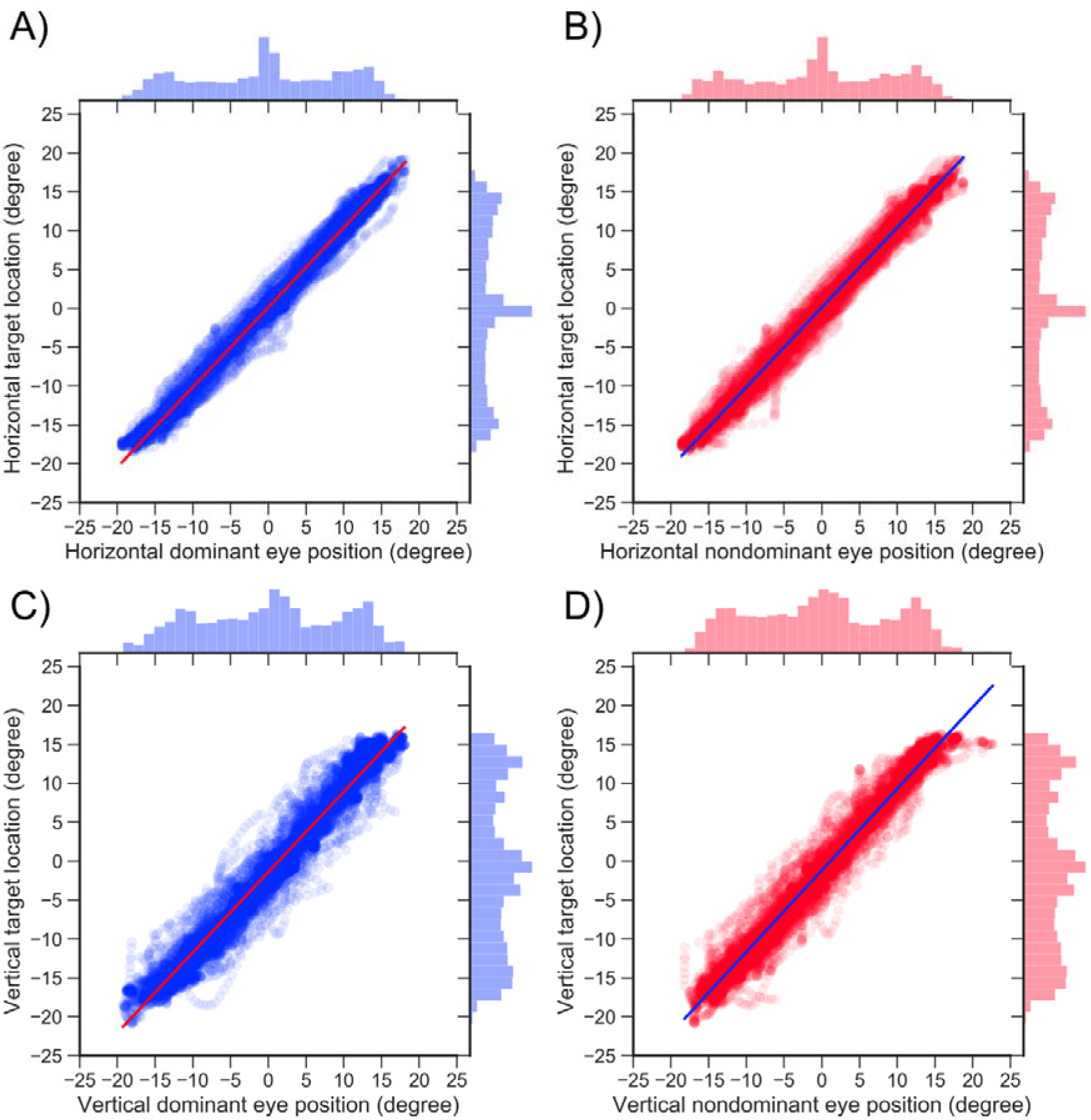
Correlations between horizontal (A, B) and vertical (C, D) target locations and eye positions. The blue and red dots indicate the relationships between the target and the dominant or nondominant eye velocity. The red and blue lines indicate the regression lines. The histograms at the upper and right sides indicate the distribution between the target location and the dominant or nondominant eye position.

The latencies of the horizontal and vertical SPEM were 97.3 ± 26.9 and 115 ± 34.1 ms for the dominant eye (*P* = 0.140), respectively. The latencies of horizontal and vertical SPEM for the nondominant eye were 108.2 ± 28.6 and 119.1 ± 33.9 ms (*P* = 0.31), respectively. The latencies of horizontal SPEM were significantly and positively correlated with the latencies of vertical SPEM in the dominant (*adjusted R*^*2*^ = 0.824, *P* < 0.001) and nondominant (*adjusted R*^*2*^ = 0.661, *P* = 0.002) eyes.

## Discussion

Our results show that the combination of VOG and SSD can be used to evaluate the SPEM of nine directions, and this method can be translated into clinical settings without changing the testing methods. The SSD recognized the target with high accuracy, and the target location was significantly and positively correlated with the positions of both eyes in main study (Figs. 5 and 6). The small variation in values in the pilot study (Figs. 4 and 6) may be attributable to the fact that the examiner moved the target by hand, making the SSD’s bounding box more susceptible to deformation because the center coordinates of the bounding box were calculated and converted from pixels to degrees. Nevertheless, the average precision of the SSD on one class of targets was 97.7% in the test dataset. We considered that SSD has sufficient accuracy and can be accumulated to evaluate SPEM.

Latency or reaction time of SPEM is one of the parameters required to evaluate strbmismus.^18, 21, 32^ In our cohort, the mean latencies of horizontal SPEM were consistent with those reported by Erkelens and Engel.^33, 34^ The latencies of horizontal and vertical SPEM were not significantly different. Although the testing methods were different, our findings support those of Rottach et al. who reported that the latencies of the horizontal and vertical SPEM were not significantly different.^35^ These findings suggest that the current system can accurately determine SPEM in healthy individuals. Therefore, we will investigate subjects with abnormal SPEM in further studies.

A limitation of our system is the low sampling rate due to the use of a scene camera. If the target moves at a high speed, the target captured by the scene camera is blurred, and the accuracy of SSD decreases. Therefore, our system cannot accurately evaluate saccadic eye movement. In future research, we plan to update our system to improve the sampling rate of the scene camera to accurately analyze saccadic eye movement.

## Conclusions

SSD recognized the target that was moved manually with high accuracy, and the target location was significantly and positively correlated with the positions of both eyes that were measured by VOG. Our findings suggest that the combination of VOG and SSD is suitable in evaluating SPEM in the clinic.

## Data Availability

The data that support the findings of this study are available from the corresponding author, [MH], upon reasonable request.

## Acknowledgments

All authors thank Mika Suda for illustrating the mascot character. This work was supported by Grands-in-Aid for Early-Career Scientists, Scientific Research (A) and (B), Challenging Exploratory Research, Japan Society for the Promotion of Science, 19K20728 (MH), 18H04116 (MH), 20K04271 (MH), 19K21783 (MH), and Charitable Trust Fund for Ophthalmic Research in Commemoration of Santen Pharmaceutical’s Founder (MH).

## Author contributions

MH conceived the project and designed the experiments. MH produced the apparatus. MH performed experiments. MH, TH,, YI, EW, and AM analyzed the data. MH, TH, and AM wrote the manuscript. All authors reviewed the manuscript.

